# Knowledge, attitudes and practices of Italian general practitioners on dietary supplements

**DOI:** 10.64898/2026.03.11.26348116

**Authors:** Laura Brunelli, Marco Driutti, Luca Arnoldo, Stefano Celotto, Federica Cozzarin, Elisa Mansutti, Maria Parpinel

## Abstract

**Objective:** The aim of the study was to investigate knowledge, attitudes and practices (KAP) of Italian general practitioners regarding the use of food supplements.

**Subject and Methods:** A 62-question survey, adapted from a tool used among pharmacists, was sent to Italian general practitioners. The survey contains two sections: A) KAP questions (28 true/false, 34 on a 5-point Likert scale) and B) socio-demographic questions.

**Results:** 233 questionnaires were collected from March to July 2024. 44.6% of responders were male, 54.9% female. The majority (68.2%) came from an urban environment and 53.65% worked with other colleagues. Most of the general practitioners (69.1%) showed an adequate knowledge of the subject (> 60% of correct answers). The overall reliability of the test was rated as acceptable (alfa*>*0.7). 98.7% of general practitioners agreed that they have an important role in prescribing dietary supplements, and 98.3% are involved in lifestyles counselling, but only 66.52% felt that they had adequate training on this subject.

**Conclusion:** General practitioners recognize their pivotal role in supplement prescription, but, even if their knowledge scores were adequate, they feel a need of more specific education on the topic.

## Introduction

The Regulation 178/2002 of the European Community (EC) and the Italian Legislative Decree 169/2004 define food supplements as dietary products that supplement the diet [1] and do not have therapeutic properties or prevent / cure diseases [2]. In Italy, supplements can be sold without a prescription in various authorized outlets (e.g. pharmacies, para-pharmacies, herbal stores) and authorized e-commerce platforms under the regulations of Legislative Decree 114/1998 [3].

According to a recent study by CENSIS (Social Investment Study Center), 32 million Italian citizens use food supplements and vitamins, with a 12% increase in sales since 2020 related to the COVID-19 pandemic. The increase in online sales facilitates access to these products from unauthorized sources and without professional advice, making it difficult to estimate real utilization. In 2023, 230 RASFF (Rapid Alert System For Food and Feed) notifications (7.2%) for non-compliant products were related to food supplements, often due to mislabeling or false health claims [4].

Serious health problems may be caused by supplement misuse and abuse such as allergic reactions, drug interactions [5], toxicity and even cancer [6–10].

General practitioners (GPs) are central to managing these risks, yet few studies investigate their knowledge on this topic [11]. Building on our previous work with pharmacists [12], this study investigates the knowledge, attitudes, and practices of Italian GPs regarding food supplements.

## Methods

### Primary Aim

To assess the level of knowledge and describe the attitudes and prescribing behaviors of Italian general practitioners regarding the use of food supplements.

### Secondary Aim

To assess the differences in knowledge, attitudes and prescribing behaviors of general practitioners regarding the use of food supplements by subgroups in terms of age, gender, professional experience, and presence or absence of specific training on the subject.

### Study design

Cross-sectional observational study.

### Setting

The population of general practitioners at national level consists of 35,398 physicians according to the data of the Italian Institute of Statistics (ISTAT) for the year 2024 [4]. Italian general practitioners were invited to participate in the study by e-mail from March 2024 to July 2024. The general practitioners were informed by the Scientific Society of Italian General Practitioners (SIMG) about the survey, the reasons for the study and its objectives. Reminder e-mails were sent during the survey period to increase the response rate. Participation was voluntary and was not remunerated. Data were stored in anonimous form on EU survey platform for the period of the study and analyzed in the month of september 2024 by the researchers, no data were shared outside of the research team. The study protocol was approved by the Institutional Review Board of the University of Udine, Italy.

### Eligibility criteria for participation

Activity as a general practitioner on Italian territory willingness to participate in the survey.

### Exclusion criteria

*Retirement*

### Instrument

The questionnaire used in the study was developed by the research team based on the scientific literature and previous experience gained in the study of the same subject among pharmacists in the Friuli Venezia Giulia region (Italy) [12]. The questions were adapted in relation to the Italian regulatory context and in relation to the sale of food supplements, as well as to the target population of the study. The questionnaire, available both in Italian (the original language) and in English translation in the supplementary materials, consists of two main sections: KAP section: a total of 62 questions. Questions (Q) 1 to 28 relate to knowledge and are asked as true/false; questions 29 to 62 relate to attitudes, opinions and behaviors and are expressed as agree/disagree on a 5-point Likert scale, with 1 indicating complete disagreement and 5 indicating complete agreement. This part of the survey is ide- ally divided into five groups of questions representing five latent variables: knowledge of the subject, need for continuing education, perception of the role of the general practitioner, perception of external influences, counselling behavior. Demographic section: nine multiple-choice questions useful to stratify the answers given by general practitioners according to demographic data. The questionnaire refers only to vitamin supplements and nutritional supplements for oncohaematologic patients.

### Dependent variables

Individual knowledge, attitudes, and practices were measured as dependent variables based on questionnaire responses. General knowledge was tested through true*/* false questions (KAP section), awarding one point for each correct answer. A score above 60% indicated adequate knowledge. general practitioners’ attitudes and practices were evaluated using a 5-point Likert scale, focusing on four sub-variables: the need for continuing education, the general practitioners’ role, external influences, and counseling behavior. Participants who scored above the group median were considered in agreement. The degree of agreement and mean scores were analyzed to identify differences between subgroups within the population.

### Independent variables

Age, gender, years of experience as a general practitioner, a general practitioner specific training, presence of specific training and organizational context were considered as independent variables. All of these variables were categorized as categorical.

### Sample size

The survey was sent to a total of 7952 of general practitioners and 233 complete responses were collected, corresponding to a response rate of 2.9% Under the null hypothesis that 50% of the above population of 35,398 active general practitioners in Italy in 2024 have adequate knowledge of the subject (more than 60% correct answers), the sample size achieved was sufficient to calculate this proportion with a margin of error of 6.4% at a 95% confidence level under the normal distribution hypothesis.

### Statistical analysis

Descriptive analyses were performed for the entire sample and the following subgroups: gender (where gender refers to the socially constructed roles, behaviors and identities of female, male and gender-diverse people), age group, years of professional experience, presence of specific training, organizational context.

The categorical variables were summarized as absolute numbers and relative frequencies. For the scored variables (total knowledge score), mean, median and standard deviation were calculated and the Shapiro-Wilk test was used to test for normal distribution.

Differences in knowledge scores between subgroups were tested using Mann Whitney and Kruskall-Wallis tests. The existence of a correlation between the presence of adequate knowledge (more than 60% correct answers) and the socio-demographic groups was tested using the Chi Square independence test; if more than one category was present, a reference category was chosen to test independence. For Likert scales, the degree of agreement between participants was tested by comparing participants’ responses to the indifference threshold of 3 out of 5 on the Likert scale (a score of 4 or 5 was considered agreement with the statement). The mean scores for each question were also compared between the different subgroups using Mann-Whitney and Kruskall-Wallis tests. The reliability of each part of the survey (need for continuing education, perception of the role of the general practitioner, perception of external influences, counselling behavior) was tested using Cronbach’s Alpha and Mc Donald’s Omega.

A statistical significance level of 0.05 was selected for the statistical test, with the null hypothesis that there are no differences between the groups. All statistical analyses were performed with SAS Enterprise Guide, version 7.1.

## Results

Overall, a total of 233 questionnaires were collected. 128 of the participants identified themselves as female (54.9 %), 104 as male (44.6 %) and 1 as non-binary. 144 respondents (61.8 %) had specific training in family medicine. The majority (124; 53.2 %) had more than 10 years of professional experience, 34 (14.6%) between six and 10 years, 70 (30.0 %) 1-5 years, and 5 (2.15 %) less than one year. Most of the general practitioners surveyed (187, 80.25 %) work with other doctors, 125 of them in group medicine with other general practitioners in the same clinic and 62 in a general practitioner professional network (Table 1).

**Table 1:** Population description.

| Characteristic | Frequency | Percent (%) |
| --- | --- | --- |
| **Gender** |  |  |
| Female | 128 | 54.9 |
| Male | 104 | 44.6 |
| Non-binary | 1 | 0.4 |
| Age (years) | **Mean (SD)**<br>46.1 (14.1) | **CI 95%**<br>[44.3-47.9] |
| **GP specific formation** |  |  |
| No | 89 | 38.2 |
| Yes | 144 | 61.8 |
| **Professional experience (years)** |  |  |
| <1 | 5 | 2.15 |
| 1-5 | 70 | 30.0 |
| 6-10 | 34 | 14.6 |
| 11-15 | 32 | 17.7 |
| 16-20 | 8 | 3.4 |
| >20 | 84 | 36.1 |
| **Specific formation on dietary supplements** |  |  |
| No | 192 | 82.4 |
| Yes | 41 | 17.6 |
| **Work organization** |  |  |
| Group medicine | 125 | 53.7 |
| GP professional network | 62 | 26.6 |
| Working alone | 46 | 19.7 |
| **Working in countryside?** |  |  |
| Yes | 74 | 31.8 |
| No | 159 | 68.2 |

### Knowledge about dietary supplements

69.1% of general practitioners answered at least 60% of the questions correctly, with a mean score of 18.9 out of 28 (95% CI 18.6-19.3). No significant differences were observed between subgroups, except between rural and urban areas (Table 2). Table 3 summarizes the results for each question: fewer than half of the 12 questions had a correct response rate above 75%. The highest-performing question concerned the link between high-dose protein supplements and kidney damage (226/233 correct answers). However, many respondents confused dietary supplements with pharmacological products, with only 17.6% answering correctly. Knowledge of multivitamin supplements was generally poor, particularly regarding calcium and vitamin D (28.8% correct answers) and polyunsaturated fatty acids (33.0% correct answers).

**Table 2:**
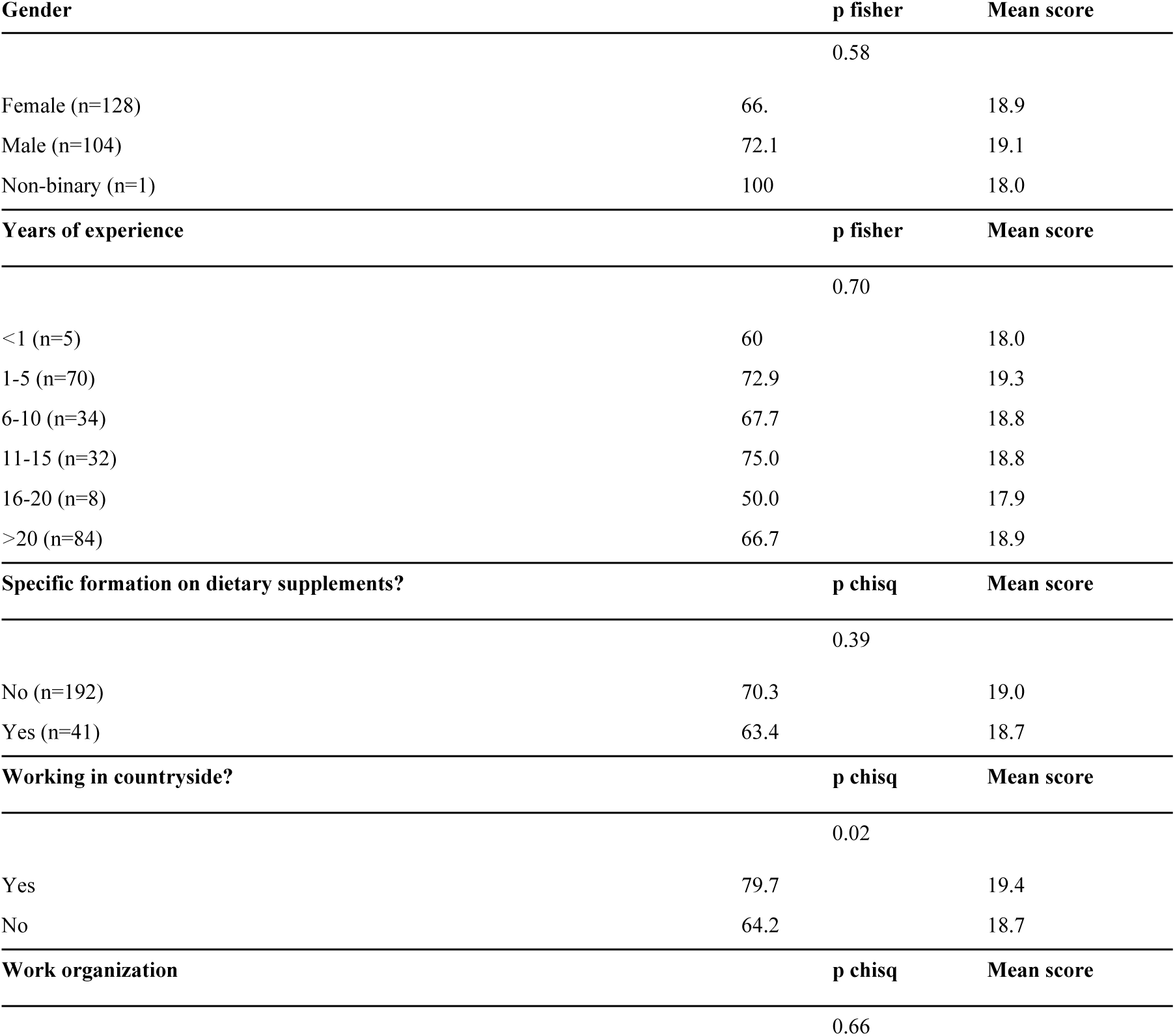
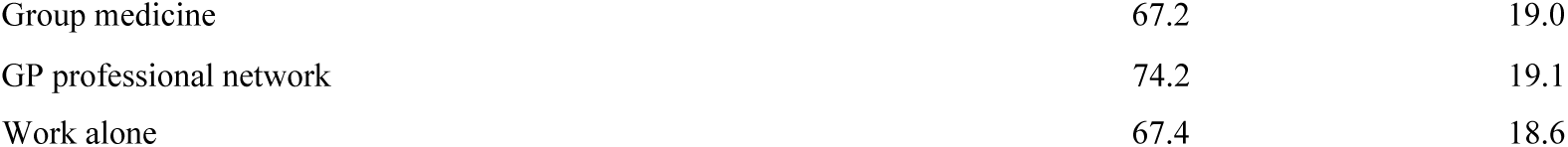
Sub-group analysis of GPs’ knowledge about dietary supplements. The results include gender, years of experience, specific formation, and work setting.

**Table 3:** GPs’ Knowledge about dietary supplements.

| Question code | Question | Overall knowledge <sup>a</sup> |
| --- | --- | --- |
| Q1 | A balanced provision of nutrients is guaranteed by the adoption of a varied diet rather than by using multivitamin supplements | 66.5 |
| Q2 | Dietary supplements contain only substances that can be found in food | 17.6 |
| Q3 | Multivitamin supplements may contain non-labelled toxic ingredients | 39.5 |
| Q4 | Increasing evidence supports the hypothesis that some types of cancer are caused by the abuse of multivitamins (commonly used as antioxidants) | 48.5 |
| Q5 | Some lifestyles can reduce the absorption of vitamins or cause their complete depletion | 96.6 |
| Q6 | The chronic use of some drugs can cause a significant deficit of vitamins | 97.0 |
| Q7 | Important drug interactions and side effects can be enhanced by the concomitant intake of vitamin supplements | 85.4 |
| Q8 | Excessive intake of Vitamin D (higher than the RDA) can cause loss of appetite, vomiting and increased urination frequency | 79.4 |
| Q9 | The recommended daily allowance (RDA) for folic acid in a man adult is 400 $\mu$ grams | 71.2 |
| Q10 | The recommended daily allowance (RDA) for folic acid in women increases during both pregnancy and lactation | 89.3 |
| Q11 | The recommended daily allowance (RDA) in men and women between 51 and 70 years for Vitamin D is 400 IU | 53.7 |
| Q12 | Excessive intake of Vitamin E can increase the risk of hemorrhagic stroke, development of skin hematomas, bleeding and headache | 49.8 |
| Q13 | Different formulations affect iron absorption | 95.3 |
| Q14 | The presence of cracks in the corners of the mouth may indicate a deficit of Vitamin B12 | 88.0 |
| Q15 | The presence of dandruff may indicate a Biotin deficiency | 63.5 |
| Q16 | Conjunctival dryness can indicate a Vitamin A deficiency | 70.8 |
| Q17 | The intake of high doses of antioxidant vitamins (A, C, E) can interfere with the effectiveness of some chemotherapy drugs | 75.5 |
| Q18 | The administration of branchedchain amino acids and acid eicosapentenoic (EPA) can help reducing loss of weight and muscle mass in cancer patients | 55.8 |
| Q19 | Pharmacists can dispense all vitamin supplements without prescription | 63.1 |
| Q20 | Vitamin D supplementation reduces cardiovascular risk | 55.8 |
| Q21 | Calcium and vitamin D supplementation in the general geriatric population is an effective and safe way to reduce the risk of fractures and osteoporosis | 28.8 |
| Q22 | Vitamin C supplementation during the flu season reduces the risk of becoming ill with respiratory diseases | 51.1 |
| Q23 | Supplementation of polyunsaturated fatty acids and omega three has proven efficacy in reducing the incidence of cardiovascular events | 33.1 |
| Q24 | The use of protein dietary supplements may be associated with eating disorders | 67.4 |
| Q25 | The use of protein supplements may be helpful in reducing muscle mass and weight loss in the healthy elderly | 82.0 |
| Q26 | High-dose protein supplementation may cause kidney damage | 97.4 |
| Q27 | If there is no risk to the patient, it is permissible to administer a supplement even in the absence of evidence of proven efficacy | 82.0 |
| Q28 | Any adverse effects (related to taking the supplement and reported by the patient) should be reported to the appropriate Health Authority | 91.0 |

<sup>a</sup> The overall knowledge was expressed as % of questionnaires reporting the correct answer.

### Attitudes and practices towards dietary supplements

Table 4 summarizes the mean Likert-scale for each question, stratified by respondents’ main characteristics and minimum acceptable level of knowledge (more than 16 correct answers).

**Table 4:** Attitude and Practices Analysis. Responses on a Likert scale by groups.

| Sub-domain | Question | ≤ 10 Years | >10 Years | No Training | Yes Training | ≤16 Correct | >16 Correct |
| --- | --- | --- | --- | --- | --- | --- | --- |
| **Need for Continuing Education (7questions)** |  |  |  |  |  |  |  |
| Q29 | I have sufficient information regarding the indications for the specific use of supplements and dietary supplements for my patients. | 2.9 | 2.8 | 3.0 | 2.9 | 3.1 | 2.9 |
| Q30 | I have sufficient information regarding the adverse effects of vitamins when taken in dosages above the recommended daily dose. | 2.9 | 2.6* | 3.1* | 2.8 | 3.1 | 2.8 |
| Q31 | I have sufficient information regarding contraindications of vitamin supplements in special groups of patients (e.g., diabetics, epileptics, on oral anticoagulant therapy). | 2.8 | 2.6* | 3.0* | 2.8 | 3.0 | 2.8 |
| Q32 | I have sufficient knowledge regarding the dosage and administration of vitamins as dietary supplements. | 3.0 | 3.0 | 3.1 | 3.0 | 3.2 | 2.9* |
| Q33 | I have sufficient knowledge regarding drug and supplement interactions. | 2.7 | 2.8 | 2.7 | 2.7 | 2.9 | 2.7 |
| Q34 | Only interprofessional and evidence-based education can ensure proper use of dietary supplements by all professionals. | 4.5 | 4.5 | 4.5 | 4.5 | 4.4 | 4.5 |
| Q35 | General Practitioners should periodically update themselves on evidence regarding vitamin supplements in print media or official/institutional websites. | 4.4 | 4.3 | 4.4 | 4.3 | 4.5 | 4.3 |
| **Role of the GP (11 questions)** |  |  |  |  |  |  |  |
| Q36 | The use of vitamins and supplements should be reported by patients to their physicians. | 4.7 | 4.6 | 4.7 | 4.7 | 4.7 | 4.7 |
| Q37 | Supplements should be considered as medications. | 3.7 | 3.7 | 3.7 | 3.7 | 3.8 | 3.8 |
| Q38 | It is advisable to ask the individual patient about his or her medical history before recommending a supplement to rule out contraindications. | 4.6 | 4.5 | 4.6 | 4.5 | 4.6 | 4.6 |
| Q39 | Dietary supplements are safe and have fewer side effects than prescription drugs for the same problem. | 2.7 | 2.7 | 2.7 | 2.7 | 2.9 | 2.6 |
| Q40 | It is the GP's responsibility to inform the patient about proper nutrition and natural sources of vitamins. | 4.3 | 4.3 | 4.3 | 4.3 | 4.4 | 4.3 |
| Q41 | GPs play a key role in supporting adequate nutrition of cancer patients, including prescribing nutritional supplements if necessary. | 4.1 | 4.0 | 4.2 | 4.1 | 4.2 | 4.1 |
| **Practices (10 questions)** |  |  |  |  |  |  |  |
| Q53 | I provide up-to-date information to my patients regarding the possible intake of supplements. | 3.6 | 3.4* | 3.7* | 3.5* | 4.1* | 3.5* |
| Q54 | I recommend supplements to all patients, being confident of their safety and effectiveness. | 2.5 | 2.4 | 2.5 | 2.3* | 3.0* | 2.3* |
| Q55 | I recommend supplements only to specific categories of patients. | 3.7 | 3.7 | 3.7 | 3.6 | 3.8 | 3.7 |
| Q56 | I happen to question the appropriateness of a supplement prescription made by others. | 3.3 | 3.3 | 3.4 | 3.3 | 3.4 | 3.4* |

The majority of surveyed general practitioners believe they play an important role in prescribing dietary supplements. Nearly all respondents (98.71%) agreed that patients should consult their doctor before taking supplements (Q36, mean score 4.7) and consider medical history (Q38, mean score 4.6). General practitioners emphasized their role in supporting the nutritional needs of older and chronically ill patients (Q41-43, mean scores 4.1-4.5), though they were neutral about the need for more education in this area (Q31-33, mean scores 2.8, 3.0, 2.7). Most confirmed active involvement in advising on lifestyle and dietary supplements (Q48-62, mean scores 4.3-4.7). Respondents indicated low external influence on supplement prescribing (Q47-51, mean scores 2.9-3.5) and stressed that prescribing should prioritize efficacy and economic evaluation (Q52, mean score 4.1). There was a neutral stance on other professionals prescribing supplements (Q56, mean score 3.3). Some differences emerged by gender and experience: women were more concerned about staying informed (Q35, mean score 4.5 vs. 4.3) and less likely to prescribe supplements without evidence of efficacy (Q54, mean score 2.6 vs. 2.3). General practitioners with over 10 years of experience demonstrated greater confidence in identifying misuse (Q30, mean score 3.1 vs. 2.6) and side effects (Q31, mean scores 3.0 vs. 2.6). The frequency of dietary supplement advice (Q53) correlated with experience, specific training, and higher knowledge scores, indicating that more experienced and better trained general practitioners were more proactive in this area. Minimal differences were found between rural and urban general practitioners, except that urban general practitioners provided advice more frequently (Q53, mean score 3.7 vs. 3.4) and displayed a more neutral attitude toward supplement use (Q54, mean score 2.6 vs. 2.2). Overall, general practitioners highlighted their role in guiding patients on supplement use while recognizing the importance of evidence-based prescribing.

### Survey reliability

The reliability of the knowledge section was quite low (*alfa* = 0.5, *omega* = 0.4), which means heterogeneity between the items, while the attitude and practice sections showed good reliability (*alfa* = 0.8, *omega* = 0.8 for the questions related to need for continuing education, *alfa* = 0.8, *omega* = 0.8 for the question on the Perception of the role of the general practitioner and *alfa* = 0.6, *omega* = 0.7 for the perception of external influences and *alfa* = 0.7, *omega* = 0.6 for Counselling behavior).

## Discussion

This study examined the knowledge, attitudes, and practices of Italian general practitioners regarding dietary supplements. While 69.1% answered over 60% of knowledge questions correctly, gaps remained in specific areas like vitamin D and omega-3 efficacy, highlighting the need for targeted training. Most general practitioners viewed themselves as key players in patient education and promoting healthy lifestyles. These attitudes were reflected in practice, with the majority actively engaging in counseling patients on dietary supplements and lifestyle choices. The findings underline the importance of supporting general practitioners with additional training to enhance their knowledge and further strengthen their role in guiding patients effectively.

A recent report by SNAMID (National Society for the Updating of General Practitioners) showed that only half of general practitioners declare to recommend food supplements to their patients [13]. The results of this study seem to confirm the trend with most of the general practitioners interviewed saying that they prescribe supplements only to specific categories of patients and for specific needs avoiding prescription on patient’s demand when not necessary. The same report shows that general practitioners most commonly prescribe food supplements to treat asthenia (53%), stress symptoms/mental fatigue (49%), inadequate nutrition in the elderly (35%) and mild anxiety symptoms (31%). Regarding prescriptions most of the general practitioners interviewed in our study say that they ask patients about food supplements frequently but the SNAMID report shows that only 30% of patients report to use food supplements during their anamnesis [14]. The SNAMID report shows that only 28% of the doctors interviewed ask their patients about supplement use and as few as 34% ask this question in relation to unexplained side effects [13]. When the general practitioners raise the topic of food supplements with patients, the reasons for prescribing a particular food supplement and how to take it are the most frequently mentioned, while the potential risks and efficacy of such products are less frequently discussed [15].

As stated before, the lack of medical supervision poses a significant risk to patient’s health and well-being both in short and in the long term. Carcinogenic effects of Vitamin A abuse and the interaction between Vitamin B6 and Parkinson’s drugs are two known examples [5; 8; 9; 10]. The results of our study highlight the concern about general practitioners’ lack of formation in this specific topic and their desire for a more specific training. Given the perception of their pivotal role in food supplement management and in patient health education, providing up to date evidence and limiting the patients’ exposure to inaccurate and harmful sources is central [15].

A recently published study of the same research group [12], investigating the knowledge and professional behavior of pharmacists in Friuli Venezia Giulia regarding food supplements, showed a good general knowledge of possible interactions between drug therapies and vitamin deficiencies, and highlighted that the pharmacist is a supportive figure for the consumer by giving the right indications on a balanced diet and natural sources of vitamins and nutrients in the diet food. However, the study found that respondents had little knowledge of the potential adverse effects of food supplements, particularly the potentially carcinogenic role associated with taking multivitamins in amounts exceeding recommended doses [8], as well as safety risks due to the possible presence of toxic ingredients not listed on the label [4].

Overall, the study found that two thirds of respondents had a low level of knowledge on this topic. Comparing our results to this study the percentage of questionnaires with more than 60% correct answers was slightly higher among general practitioners (69.1% or 161/233 vs 55.6% or 79/142) [12]. The attitudes and behaviors of the two groups of health professional, however, appear to be similar, with both placing great importance on counselling and emphasizing the need of being up to date.

As far as we know, this is the only study addressing this subject among Italian general practitioners, although some similar studies have been conducted in other countries: from Europe (Serbia [16], Portugal [17]) to middle East (Saudi Arabia [18], Egypt [19], Lebanon [20], Palestine [21]), Africa (Ethiopia [22]) and North America (United States [23] and Canada [24]). Among these studies many focus on other health professionals like pharmacists [12, 18, 19, 21, 22, 24, 25, 26], nurses [23], and dietitians [20]. A review of 44 studies found low awareness of supplement side effects and drug interactions among the general population but better knowledge among healthcare professionals, with some variation [11]. Given the widespread availability of supplements and reliance on marketing for information, healthcare professionals like doctors and dietitians play a key advisory role. Studies on general practitioners show good understanding of supplements and legal aspects, but comparisons are limited by differing questionnaires and study focuses [16, 17]. Legal and cultural differences further complicate establishing consistent knowledge standards and make difficult to give a direct comparison.

### Limitations and strengths of the study

Our study has several limitations, such as the low reliability of the knowledge question section in determining the final score. This issue may be linked to the limited number of questions in the survey investigating such a broad topic. The lack of a shared minimum knowledge base and the formation differences between professionals proved also a challenge in the questionnaire construction.

The low response rate was a problem, although it was to be expected due to the nature of online surveys. Regarding the strengths of the study, to our knowledge this is the first study aimed at general practitioners in Italy and one of the few studies available in the literature regarding these health professionals. The presence of the “attitudes and practices” section in the questionnaire provides good insight into the behaviors of the study population and sheds new light on an important topic that has rarely been addressed before.

## Conclusion

In conclusion, general practitioners’ knowledge of dietary supplements proved to be adequate on average, although only a few respondents answered more than 75% of the questions correctly. The need for further training in this area is shared by the professionals surveyed, although there were no significant differences in knowledge scores between general practitioners with and without special training in this area. This study con- firms the belief that general practitioners play an important role in patient education and participate in lifestyle counseling.

## Data Availability

All relevant data are within the manuscript and its Supporting Information files.

## Declarations

### Ethics approval and consent to participate

General practitioners gave their informed consent in writing before the questionnaire was administered. The surveys were administered through the Eu Survey platform. Emails containing the survey link were sent to eligible professionals by the main scientific association of Italian general practitioners (SIMG). Participation was voluntary and free of charge.

The study protocol was approved by the Institutional Review Board of the University of Udine, Italy. The study manuscript was written in accordance with Strobe Reporting guidelines for cross sectional studies.

### Consent for publication

Not applicable.

### Availability of data and materials

The datasets used and/or analysed during the current study are available from the corresponding author on reasonable request.

